# Knowledge and Willingness towards Human Papillomavirus Vaccination among the Parents and School Teachers of Eligible Girls in Dhaka, Bangladesh: A School-Based Cross-Sectional Study

**DOI:** 10.1101/2024.10.12.24315384

**Authors:** Samina Sultana, MD Nahid Hassan Nishan, Aklima Akter, Dalia Rahman, Fowzia Yesmin, Mohammad Delwer Hossain Hawlader

## Abstract

**Background:** Cervical cancer ranks as the common prevalent cancer, among women worldwide especially impacting low-resource countries. In Bangladesh, this accounts for 12% of all cancer cases. The development of cancer is closely linked to Human Papillomavirus (HPV) infection. Despite the availability of HPV vaccines, their uptake remains limited in Bangladesh. Thus, this research aims to assess the knowledge and willingness of parents and school teachers regarding HPV vaccination for eligible girls in Bangladesh.

**Methodology:** This study involved 406 parents and school teachers of girls aged 9-14 years from Dhaka city. A cross-sectional study design was used. Data collection was done through a questionnaire administered by interviewers after pre-testing and refinement for clarity and reliability. Analysis was carried out using Stata 17 software. Chi-square tests and logistic regression were used to uncover associations and predictors related to knowledge levels and willingness.

**Results:** Findings revealed that a majority of participants (64.04%) exhibited an understanding of HPV and cervical cancer yet a high percentage (98.28%) expressed willingness to engage in HPV vaccination initiatives. participants with primary (AOR=3.306, p<0.005), secondary (AOR=8.806, p<0.001), and higher education (AOR=5.059, p<0.001), as well as those from upper-middle-income groups (AOR=3.038, p<0.001), had significantly higher knowledge of HPV and cervical cancer.

**Conclusion:** The research emphasizes lack of knowledge regarding HPV and its vaccination among parents and educators in Bangladesh despite a willingness to vaccinate. These results emphasize the importance of tailored initiatives and better access, to health information to increase HPV vaccine acceptance and lower the incidence of cervical cancer.

## Introduction

Cervical cancer ranks as the fourth most common cancer among women globally, with a significant burden in low-resource countries [1]. In 2022, 660,000 women were diagnosed and 350,000 died from this disease worldwide [1]. In Bangladesh, cervical cancer represents 12% of female cancers, following breast cancer (19%) and esophageal cancer (11.1%) [2]. In 2020, there were 8,268 new cases and 4,971 deaths in Bangladesh [2]. Infection with HPV, a non-enveloped, double-stranded DNA virus, is a critical cause of cervical cancer [2]. Among the approximately 100 fully sequenced HPV types, high-risk types 16 and 18 are the main contributors to cervical cancer [3,4], with HPV type 16 responsible for over 50% of squamous cell carcinomas and most adenocarcinoma cases [5–8].

HPV infections typically occur in late teens or early 20s in unvaccinated populations [6,9]. The Immature metaplastic cells in the cervix, especially during puberty and early pregnancy, are highly susceptible to HPV infection [10]. However, the HPV vaccine played a major public health role in preventing HPV-related cancers [11,12]. Prophylactic vaccines, such as bivalent, quadrivalent, and nonavalent, are produced using virus-like particle (VLP) technology [11,13,14], inducing type-specific antibodies that prevent persistent HPV infection and precancerous lesions [15,16].

In 2008, the US Food and Drug Administration approved the first HPV vaccine, which prevents 70% of cervical cancers and other HPV-related diseases [17]. By 2017, 71 countries had introduced the vaccine for girls aged 9 to 13 years [18]. Bangladesh introduced the HPV vaccine in 2016, in the Gazipur district, funded by the Global Alliance for Vaccines and Immunization (GAVI) [19]. However, successful implementation requires addressing knowledge gaps and societal attitudes toward the vaccine [20].

Studies highlight the importance of evaluating vaccine-related knowledge among adolescents, parents, and school teachers. For instance, in Kampong Speu, Cambodia, 35% of women knew about cervical cancer prevention through vaccination, and 62% were willing to receive the vaccine [21]. Another study revealed high willingness among both urban and rural respondents to vaccinate themselves or their daughters [22]. These findings emphasize the influence of parents and school teachers on adolescent vaccination decisions.

Bangladesh’s National Cervical Cancer Control Program aims to integrate the HPV vaccine into the Expanded Program on Immunization (EPI) and expand cervical cancer screening and treatment [23]. The high public health burden of cervical cancer in Bangladesh makes achieving widespread vaccination essential. The WHO’s goal of vaccinating 90% of girls under 15 by 2030 further underscores this necessity [24]. However, adolescents themselves cannot make vaccination decisions; this responsibility falls to their parents, with school teachers playing a crucial role in influencing awareness and dispelling myths about vaccination [25,26].

While global studies have assessed HPV vaccine awareness, significant gaps remain, particularly in low-resource countries like Bangladesh. Therefore, this study aims to address these gaps by evaluating the knowledge and willingness of parents and school teachers towards HPV vaccination for adolescent girls in Bangladesh. This study helps to generate actionable insights that can inform future public health interventions, ultimately improving vaccine coverage and reducing cervical cancer incidence. The large sample size and focus on socio-demographic variations make this study highly relevant and generalizable.

## Materials and Methods

### Study Design

This study employed a cross-sectional design. The study involved quantitative data collection using a structured, interviewer-administered questionnaire. The structured questionnaire was designed based on a thorough review of existing literature and consultations with three experts in the field. It was pre-tested and refined to ensure clarity, reliability, and validity. **Table 1** shows that in the questionnaire, there were 8 sets of questions for knowledge and 8 sets of questions for willingness. The reliability and internal consistency of the questions were assessed through Cronbach’s alpha (α) which showed the coefficient value is higher than 0.80 indicating very strong internal consistency and reliability were present on it. Data were collected from a diverse sample of parents and school teachers from different types of schools (public, private) and various geographical locations within Dhaka (urban, and rural areas).

**Table 1:** Scale Reliability and Covariance for Knowledge and Attitude Statements.

| Scale Type | Average inter-item Covariance | Number of Items | Scale Reliability Coefficient |
| --- | --- | --- | --- |
| Knowledge Related statements | 0.0893693 | 8 | 0.8358 |
| Attitude Related statements | 0.2062145 | 8 | 0.8370 |

### Study Population and Place of Study

This study focused on parents and school teachers of adolescent girls aged 9-14 years attending primary and secondary schools in Dhaka, Bangladesh. Dhaka is the capital and largest city of Bangladesh, and more than 10 million people live here [27]. This provides a diverse demographic profile which was one of the suitable sites for conducting the study. The research was conducted across various schools within this city to ensure a representative sample reflective of the broader population.

### Study Period and Inclusion/Exclusion Criteria

The research was carried out over five months, from August 2023 to December 2023, following the approval of the research proposal. Participants included parents of adolescent girls aged 9-14 years and school teachers who directly interacted with girls within the same age range. To maintain the study’s integrity and relevance, several exclusion criteria were applied: participants residing outside Dhaka, individuals with major psychological or psychiatric conditions, parents of non-school-going adolescent girls, and individuals unwilling to provide information. These criteria ensured the inclusion of a relevant and targeted participant pool.

### Sampling Technique and Sample Size

A convenient sampling technique was employed in this study. This approach was chosen due to practical considerations such as time constraints and the accessibility of participants within the study period. Convenient sampling allowed for the efficient collection of data from readily available and willing participants, which was crucial given the logistical challenges of accessing a large and diverse population within Dhaka.

Although convenient sampling may limit the generalizability of the findings, it provided a practical solution for obtaining a sufficient sample size within the specified timeframe. Schools were selected based on their willingness to participate and their accessibility to the research team. Within these schools, parents and teachers who met the inclusion criteria and were willing to participate were approached.

The sample size was determined using a standard formula for prevalence studies:

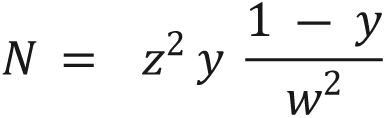

In this formula, N represents the desired sample size, Z the standard normal deviation at 5% type-1 errors (p < 0.05) with a 95% confidence interval, which is 1.96. The proportion of the population exhibiting the characteristic of interest, based on previous studies, was set at 59.2% (y = 0.592) [28]. The allowable error or precision (w) was set at 0.05. Substituting these values into the formula, the initial sample size calculated was 371.15. To account for a potential 5% dropout rate, the minimum sample size was needed to 390 participants. In this study, a total of 435 samples were collected. Upon applying exclusion criteria, removing missing values, and duplicate entries finally 406 samples were kept for the study.

### Data Collection

Data collection involved a structured, face-to-face, interviewer-administered questionnaire. Initially, the questionnaire was developed in English and then translated into Bengali to ensure comprehensibility among all participants. The development process of the questionnaire involved several steps: drafting an initial version based on a thorough literature review and expert consultations, translating the draft into Bengali by bilingual experts, pre-testing the questionnaire with a small group of parents and teachers to identify any language or comprehension issues, refining the questionnaire based on feedback from the pre-test, and conducting a field trial in two schools to finalize the questionnaire before the main data collection phase.

### Ethical Consideration

The study received approval from the Institutional Review Board (IRB) of North South University (IRB #2024/OR-NSU/IRB/0803), which was mandated by regulatory requirements. The ethical guidelines specified in the 1964 Declaration of Helsinki and its later amendments were adhered to wherever applicable. Prior to participation, all eligible respondents were provided with a detailed explanation of the study’s purpose, procedures, potential risks, and benefits. Written informed consent was obtained from each participant. The respondents were assured that their privacy and confidentiality would be strictly protected, and no part of their responses would be disclosed to unauthorized individuals under any circumstances. This commitment to ethical standards ensured the protection and respect of all participants throughout the study.

### Data analysis

The collected data were meticulously coded and entered into Stata version 17 for analysis. After applying inclusion and exclusion criteria and removal of any missing and duplicate data, the final dataset was prepared. This was checked by the supervisor as to see if the integrity and data quality were assured properly or not. Descriptive statistics were used initially to summarize the demographic characteristics of the study population, providing a clear overview of the study sample, including the distribution of demographics such as age, gender, educational level, and occupation.

For the knowledge assessment, eight specific questions were used, each with a binary response format of ‘Yes’ or ‘No.’ These questions covered key aspects of HPV and its vaccination. Participants were asked whether having multiple sexual partners is a risk factor for HPV infection, whether engaging in sexual activity at a young age increases the risk of HPV infection, whether being a smoker increases the risk of HPV infection, whether sexual contact is the primary route of transmission for HPV infection, whether the primary cause of cervical carcinoma is HPV infection, whether people can transmit HPV to their partners even without showing any symptoms, whether receiving the HPV vaccine before sexual intercourse can prevent cervical carcinoma, and whether the recommended age for receiving the HPV vaccine is nine to fourteen years old. Responses were coded as 1 for ‘Yes’ and 0 for ‘No’. A composite knowledge score was calculated for each participant, ranging from 0 to 8. The cut point was set to 4 as if the participants scoring equal and less than 4 were categorized as having poor knowledge, while those scoring higher than 4 were categorized as having good knowledge.

Willingness to vaccinate was assessed using eight Likert-scale questions, with responses scored from 1 (Strongly Disagree) to 5 (Strongly Agree). Participants were asked whether having only one sexual partner can protect an individual from HPV infection, whether HPV vaccine education should be provided to adolescents in schools, whether cervical cancer is a significant health issue for women, whether cervical cancer can cause death in women, whether men’s involvement is important in the prevention of cervical carcinoma, whether girls should receive the HPV vaccine before their first sexual intercourse, whether health information about the HPV vaccine is necessary for adolescents, and whether the HPV vaccine is effective in preventing cervical cancer. A total willingness score was calculated for each participant, ranging from 8 to 40. Participants with a total score above 24 were classified as willing to vaccinate, while those with a score of 24 or below were classified as not willing.

Bivariate analysis was performed to explore the association between demographic variables and the categorized knowledge and willingness level. Chi-square tests were used to identify significant associations between categorical variables. Variables with a p-value ≤ 0.2 in the bivariate analysis were included in the logistic regression model to ensure that potentially relevant factors were not excluded prematurely.

To adjust for potential confounders and to determine the independent effect of each variable on the outcomes of interest, multivariate logistic regression was employed. In this step, all independent variables that showed a p-value ≤ 0.2 in the chi-square were included in the multivariate model. The final model was refined by sequentially removing non-significant variables and reassessing the fit of the model. The significance level was shown as the level of strength as _Ψ_ for a p-value less than 0.05, _ΨΨ_ for a *P-value* less than 0.01, and _ΨΨΨ_ for a p-value less than 0.001. Adjusted odds ratios (AOR) with 95% confidence intervals (CI) were also shown to identify significant predictors. The results of the analysis were presented using text, tables, and figures to provide a comprehensive depiction of the findings.

## Results

In this study, a total of 406 respondents were included encompassing a diverse population in terms of age, participant type, residence, education level, and socioeconomic status. **Table 2** shows the demographic characteristics of the participants showing that the majority were aged between 31-40 years (52.96%). Most respondents were parents (69.21%), while the rest were school teachers (30.79%). The sample was nearly evenly split between urban (49.51%) and rural (50.49%) residents. Education levels varied, with higher education being the most common (47.78%) among the participants. Socioeconomic status was also balanced, with 50.25% of participants classified as lower middle income and 49.75% as upper middle income.

**Table 2:** Demographic characteristics of the participants.

| Characteristics | Frequency (N) | Percentage (%) |
| --- | --- | --- |
| **Age group** |  |  |
| 20-30 y | 65 | 16.01 |
| 31-40 y | 215 | 52.96 |
| 41-50 y | 114 | 28.08 |
| Above 50 y | 12 | 2.96 |
| **Participant type** |  |  |
| Parents | 281 | 69.21 |
| School Teacher | 125 | 30.79 |
| **Resident** |  |  |
| Urban | 201 | 49.51 |
| Rural | 205 | 50.49 |
| **Education Level** |  |  |
| No Education | 55 | 13.55 |
| Primary | 64 | 15.76 |
| Secondary | 93 | 22.91 |
| Higher | 194 | 47.78 |
| **Socio Economic Status** |  |  |
| Lower Middle Income | 204 | 50.25 |
| Upper Middle Income | 202 | 49.75 |

**Table 3** provides a detailed breakdown of the responses to knowledge and willingness-related statements. There were a significant proportion of respondents (60.84%) recognized that having multiple sexual partners increases the risk of HPV infection, and (55.17%) understood that receiving the HPV vaccine before sexual intercourse can prevent cervical carcinoma. Additionally, a substantial majority (87.37%) believed that providing HPV vaccine education in schools is essential, and (83.98%) agreed that men’s involvement is important for the prevention of cervical carcinoma.

**Table 3:** Distribution of Knowledge and Willingness Related Statement Among Respondents.

| Statements | Freq. | Percent |
| --- | --- | --- |
| **Knowledge Related** |  |  |
| **Multiple sexual partners increase HPV infection risk** |  |  |
| No | 159 | 39.16 |
| Yes | 247 | 60.84 |
| **Early sexual activity increases HPV infection risk** |  |  |
| No | 330 | 81.28 |
| Yes | 76 | 18.72 |
| **Smoking increases HPV infection risk** |  |  |
| No | 202 | 49.75 |
| Yes | 204 | 50.25 |
| **Sexual contact is the primary HPV transmission route** |  |  |
| No | 219 | 53.94 |
| Yes | 187 | 46.06 |
| **HPV infection is the primary cause of cervical carcinoma** |  |  |
| No | 260 | 64.04 |
| Yes | 146 | 35.96 |
| **HPV can be transmitted without symptoms** |  |  |
| No | 269 | 66.26 |
| Yes | 137 | 33.74 |
| **HPV vaccine before sex prevents cervical carcinoma** |  |  |
| No | 182 | 44.83 |
| Yes | 224 | 55.17 |
| **The recommended age for the HPV vaccine is 9-14 years** |  |  |
| No | 235 | 57.88 |
| Yes | 171 | 42.12 |
| **Willingness Related** |  |  |
| **I believe having one sexual partner protects from HPV** |  |  |
| Strongly Disagree | 10 | 2.46 |
| Disagree | 14 | 3.45 |
| Neutral | 56 | 13.79 |
| Agree | 190 | 46.80 |
| Strongly Agree | 136 | 33.50 |
| **I support providing HPV vaccine education in schools** |  |  |
| Strongly Disagree | 1 | 0.25 |
| Disagree | 11 | 2.71 |
| Neutral | 19 | 4.68 |
| Agree | 133 | 32.76 |
| Strongly Agree | 242 | 59.61 |
| **I consider cervical cancer a significant issue for women** |  |  |
| Strongly Disagree | 1 | 0.25 |
| Disagree | 2 | 0.49 |
| Neutral | 14 | 3.45 |
| Agree | 140 | 34.48 |
| Strongly Agree | 249 | 61.33 |
| **I believe cervical cancer can cause death** |  |  |
| Strongly Disagree | 2 | 0.49 |
| Disagree | 5 | 1.23 |
| Neutral | 10 | 2.46 |
| Agree | 154 | 37.93 |
| Strongly Agree | 235 | 57.88 |
| **I think men's involvement is important for prevention** |  |  |
| Strongly Disagree | 1 | 0.25 |
| Disagree | 10 | 2.46 |
| Neutral | 43 | 10.59 |
| Agree | 169 | 41.63 |
| Strongly Agree | 183 | 45.07 |
| **support girls getting the HPV vaccine before their first sex** |  |  |
| Strongly Disagree | 2 | 0.49 |
| Disagree | 7 | 1.72 |
| Neutral | 48 | 11.82 |
| Agree | 199 | 49.01 |
| Strongly Agree | 150 | 36.95 |
| **I believe HPV vaccine information is necessary for adolescents** |  |  |
| Strongly Disagree | 0 | 0.00 |
| Disagree | 4 | 0.99 |
| Neutral | 10 | 2.46 |
| Agree | 187 | 46.06 |
| Strongly Agree | 205 | 50.49 |
| **I agree that the HPV vaccine prevents cervical cancer** |  |  |
| Strongly Disagree | 3 | 0.74 |
| Disagree | 5 | 1.23 |
| Neutral | 32 | 7.88 |
| Agree | 196 | 48.28 |
| Strongly Agree | 170 | 41.87 |

Among the participants, a majority (72.17%) had heard about cervical cancer, and (88.42%) had no history of cervical cancer in their family. Most respondents (92.12%) indicated no history of sexual activity among their girls, and (67.00%) did not express fear of sexual transmission of HPV. Regarding HPV vaccination, (29.56%) had received the vaccine. These findings highlight a high level of awareness about cervical cancer and a substantial portion of participants taking preventive measures such as HPV vaccination.

**Table 4:** Reproductive Health Factors vs. Human Papillomavirus Vaccine Awareness Among Participants.

| Characteristics | Response | Frequency | Percent |
| --- | --- | --- | --- |
| Heard about Cervical Cancer | Yes | 293 | 72.17 |
|  | No | 113 | 27.83 |
| History of Cervical Cancer | No | 359 | 88.42 |
|  | Yes | 47 | 11.58 |
| History of Sexual Activity | No | 374 | 92.12 |
|  | Yes | 32 | 7.88 |
| Fear of Sexual Transmission | No | 272 | 67.00 |
|  | Yes | 134 | 33.00 |
| Taken HPV Vaccine | No | 286 | 70.44 |
|  | Yes | 120 | 29.56 |

After properly scoring and labeling the knowledge and willingness category the results indicated that the majority of participants had poor knowledge about the HPV vaccine and cervical cancer, with (64.04%) demonstrating poor knowledge and only (35.96%) showing good knowledge. However, the willingness to participate in HPV vaccination programs was notably high, with (98.28%) of participants expressing their willingness.

**Figure 1.**
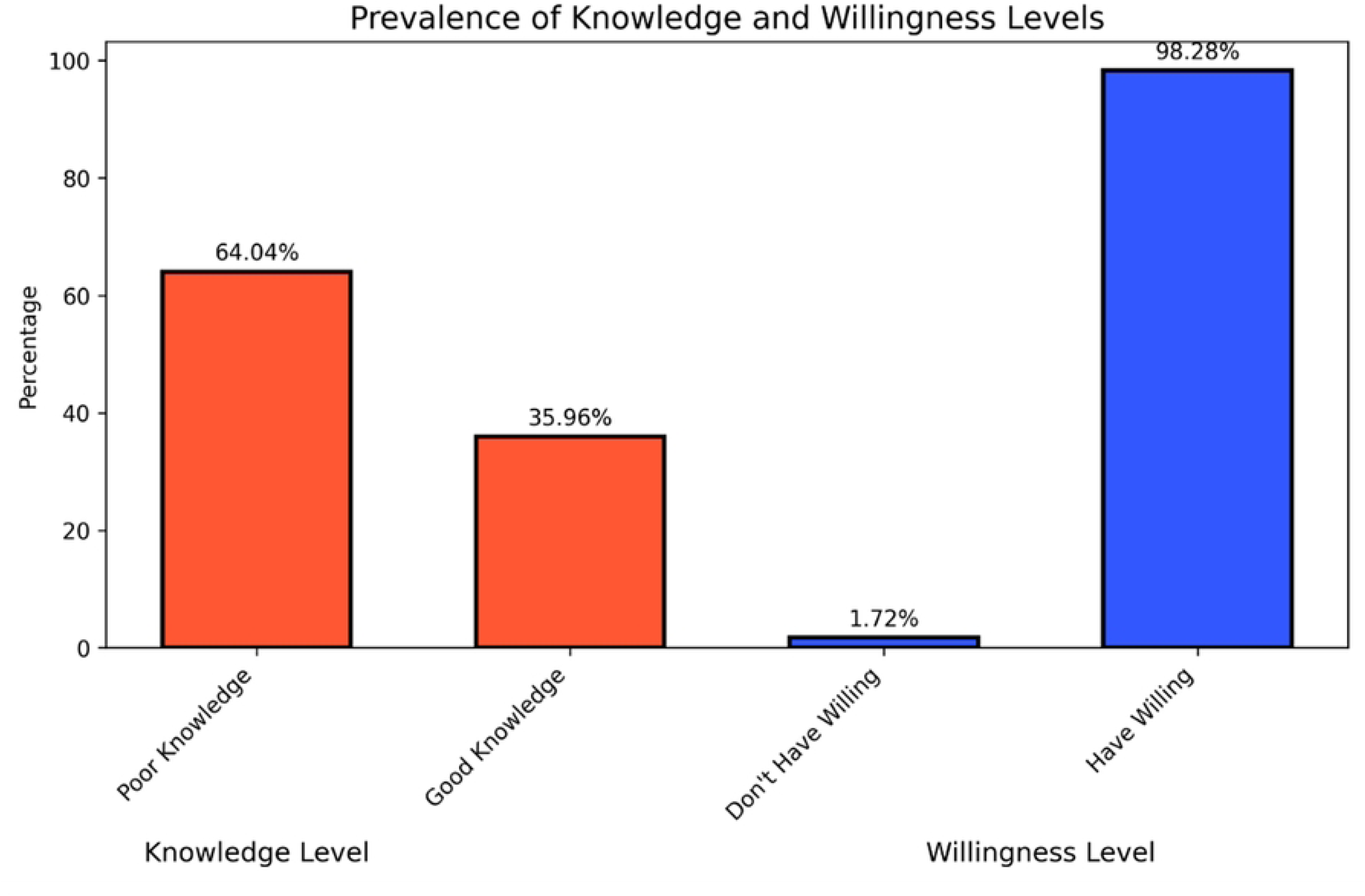

Bar chart showing the prevalence (%) of knowledge and Willingness level according to their category.

The association analysis revealed significant relationships between knowledge levels and various demographic characteristics. The age group was significantly associated with knowledge levels (X²=10.721, p=0.013), with the highest knowledge observed in the 31-40 years age group. Participant type also showed a significant association (X²=7.287, p=0.007). Residence was another significant factor (X²=18.367, p<0.001), with urban residents having higher knowledge levels than rural residents. Moreover, Education level (X²=37.068, p<0.001) and socioeconomic status (X²=42.073, p<0.001) were both significantly associated with knowledge levels, highlighting that higher education and better socioeconomic status correlate with better knowledge about HPV vaccine and cervical cancer.

However, willingness to participate in HPV vaccination programs didn’t show any significant association.

**Table 5:** Association between Knowledge and Willingness Levels across Demographic Characteristics.

| Characteristics | Good Knowledge<br>N (%) | X<sup>2</sup> Value | P-value | Have Willingness<br>N (%) | X<sup>2</sup> Value | P-value |
| --- | --- | --- | --- | --- | --- | --- |
| **Age Group** |  | 10.721 | 0.013<sup>Ψ</sup> |  | 4.461 | 0.216 |
| 20-30 y | 15 (10.27) |  |  | 64 (16.04) |  |  |
| 31-40 y | 73 (50.00) |  |  | 213 (53.38) |  |  |
| 41-50 y | 53 (36.30) |  |  | 111 (27.82) |  |  |
| Above 50 y | 5 (3.42) |  |  | 11 (2.76) |  |  |
| **Participant Type** |  | 7.287 | 0.007<sup>ΨΨ</sup> |  | 0.016 | 0.898 |
| Parents | 89 (60.96) |  |  | 276 (69.17) |  |  |
| School Teacher | 57 (39.04) |  |  | 123 (30.60) |  |  |
| **Residence** |  | 18.367 | <0.001<sup>ΨΨΨ</sup> |  | 1.369 | 0.242 |
| Urban | 93 (63.70) |  |  | 196 (49.12) |  |  |
| Rural | 53 (36.30) |  |  | 203 (50.88) |  |  |
| **Education Level** |  | 37.068 | <0.001<sup>ΨΨΨ</sup> |  | 2.959 | 0.398 |
| No Education | 4 (2.74) |  |  | 53 (13.28) |  |  |
| Primary | 13 (8.90) |  |  | 63 (15.79) |  |  |
| Secondary | 39 (26.71) |  |  | 93 (23.31) |  |  |
| Higher | 90 (61.64) |  |  | 190 (47.62) |  |  |
| **Socioeconomic Status** |  | 42.073 | <0.001<sup>ΨΨΨ</sup> |  | 1.338 | 0.247 |
| Lower Middle Income | 42 (28.77) |  |  | 202 (50.63) |  |  |
| Upper Middle Income | 104 (71.23) |  |  | 197 (49.37) |  |  |

Denote: (Chi<sup>2</sup> significance: <sup>Ψ</sup> = p-value<0.05, <sup>ΨΨ</sup> = p-value=<0.01, <sup>ΨΨΨ</sup> = p-value<0.001)

Through the logistic regression model, the study analyzed the demographic factors influencing knowledge levels about HPV and cervical cancer. Significant findings indicated that participants with primary education (AOR=3.306, p<0.005), secondary education (AOR=8.806, p<0.001) and higher education (AOR=5.059, p<0.001) had considerably higher likelihood of knowledge levels compared to those with no education. Additionally, individuals from upper-middle-income groups were significantly more knowledgeable than those from lower-middle-income groups (AOR=3.038, p<0.001). These results highlight the critical role of higher education and better socioeconomic status in enhancing awareness and knowledge about HPV and cervical cancer.

**Table 6:** Demographic factors associated with the Knowledge level.

| Characteristics | AOR | St. Error | [95% Conf. Interval] |
| --- | --- | --- | --- |
| **Age Group** |  |  |  |
| 20-30 y | Ref |  |  |
| 31-40 y | 1.598 | 0.602 | 0.763 - 3.346 |
| 41-50 y | 1.615 | 0.649 | 0.735 - 3.549 |
| Above 50 y | 2.548 | 1.883 | 0.598 - 10.847 |
| **Participant Type** |  |  |  |
| *Parents* | Ref |  |  |
| *School Teachers* | 1.278 | 0.396 | 0.697 - 2.344 |
| ***Residence*** |  |  |  |
| *Urban* | Ref |  |  |
| *Rural* | 0.674 | 0.180 | 0.400 - 1.138 |
| ***Education Level*** |  |  |  |
| *No Education* | Ref |  |  |
| *Primary* | 3.306* | 2.032 | 0.991 - 11.030 |
| *Secondary* | 8.806*** | 5.186 | 2.776 - 27.928 |
| *Higher* | 5.059*** | 3.082 | 1.533 - 16.698 |
| ***Monthly Income*** |  |  |  |
| *Lower Middle Income* | Ref |  |  |
| *Upper Middle Income* | 3.038*** | 0.838 | 1.769 - 5.217 |

Denote: (P-value indication: \* = P-value<0.05, \*\* = P-value=<0.01, \*\*\* = P-value<0.001, AOR=adjusted odds ratio)

## Discussion

The results of this study highlight several important aspects regarding the knowledge and willingness of parents and school teachers of adolescent girls toward the HPV vaccination in Bangladesh. The demographic characteristics revealed that the majority of participants were aged between 31-40 years, with a significant proportion being parents rather than school teachers. This demographic distribution is important as it suggests that the primary caregivers, who are directly responsible for the health decisions of adolescent girls, form the bulk of the respondents.

The knowledge-related findings showed that a significant portion of respondents recognized that having multiple sexual partners increases the risk of HPV infection and that receiving the HPV vaccine before sexual intercourse can prevent cervical carcinoma. However, there were notable gaps in knowledge, such as the misunderstanding that HPV infection is not the primary cause of cervical carcinoma and the lack of awareness that HPV can be transmitted without symptoms. These findings align with previous studies where awareness about HPV and its link to cervical cancer is generally low. For instance, similar gaps in knowledge, suggest that the lack of comprehensive sexual health education and cultural taboos surrounding discussions of sexual health contribute to these misconceptions [29,30].

Furthermore, the study found that urban residents and those with higher education levels had significantly better knowledge about HPV and cervical cancer. Urban residents typically have better access to health information and services, which likely contributes to their higher knowledge levels [31]. Additionally, higher education is often correlated with improved health literacy, enabling individuals to better understand and utilize health information [32].

Despite the gaps in knowledge, the study revealed a high willingness among respondents to participate in HPV vaccination programs. The overwhelming majority of participants expressed support for providing HPV vaccine education in schools and recognized the importance of men’s involvement in preventing cervical carcinoma. This high level of willingness can be attributed to the general recognition of cervical cancer as a significant health issue and the perceived benefits of vaccination. Previous research supports this finding, indicating that when individuals are aware of the benefits of vaccination, they are more likely to support and participate in vaccination programs [33].

The logistic regression analysis provided further insights into the key factors influencing knowledge levels about HPV and cervical cancer. Participants with primary, secondary, and higher education had a significantly higher likelihood of having better knowledge levels compared to those with no education. Additionally, individuals from upper-middle-income groups were more knowledgeable than those from lower-middle-income groups. These results emphasize the critical role of education and socioeconomic status in enhancing awareness and knowledge about HPV. Education equips individuals with the necessary skills to seek out, understand, and apply health information, while higher socioeconomic status provides better access to healthcare resources and information [34].

## Limitation

Although every research has limitations, one issue could be this research relies on information provided by the participants themselves which could lead to biases like wanting to appear as memory-related biases. People might exaggerate behaviors that are seen positively or downplay those that are not affecting the accuracy of the results. Moreover, because the study only captures a moment in time through its cross-sectional design it can’t fully establish cause-and-effect relationships or track changes over time. Additionally, although the research involves individuals from both urban and rural areas using a convenience sampling method might limit how broadly we can apply the findings to the population. Finally, even though steps were taken to make sure the survey was reliable and valid difference, in culture and language might still influence how participants interpret questions and provide responses potentially impacting what conclusions we draw from the study.

## Conclusion

The findings reveal significant knowledge gaps about HPV and its vaccination among parents and school teachers in Bangladesh, despite a strong willingness to support vaccination programs. Factors such as education level and socioeconomic status were found to significantly influence knowledge levels. The high willingness to participate in HPV vaccination programs highlights the potential for targeted educational interventions and improved vaccine access, driven by a general awareness of cervical cancer as a major health issue and the perceived benefits of vaccination. Enhancing access to accurate health information and making vaccines more available, especially in rural and lower socioeconomic areas, can substantially reduce HPV-related cervical cancer in Bangladesh. These efforts are crucial for improving the health outcomes of adolescent girls and supporting the broader public health objectives of the country.

## Data Availability

The data that support the findings of this study are available from the corresponding author, [Mohammad Delwer Hossain Hawlader], upon reasonable request.

## Abbreviation

| Abbreviation | Full Term |
| --- | --- |
| **AOR** | Adjusted Odds Ratio |
| **CI** | Confidence Interval |
| **EPI** | Expanded Program on Immunization |
| **GAVI** | Global Alliance for Vaccines and Immunization |
| **HPV** | Human Papillomavirus |
| **P-value** | Probability Value |
| **VLP** | Virus-Like Particle |
| **WHO** | World Health Organization |
| **X<sup>2</sup>** | Chi-Square |
| **$\alpha$** | Cronbach's Alpha |

## Acknowledgments

We would like to express our gratitude to all the parents and teachers for their participation. Without their support, it would be difficult to get the data.

